# Predicting Depression and Anxiety Progression in Multiple Sclerosis from Longitudinal Clinical Data Using Machine Learning

**DOI:** 10.64898/2026.06.23.26356339

**Authors:** Bernhard Specht, Samaher Garbaya, Reinhard Schneider, Ricardo Chavarriaga, Djamel Khadraoui, Zied Tayeb

## Abstract

Depression and anxiety are highly prevalent in multiple sclerosis (MS), yet tools for predicting mental health trajectories from clinical data remain limited. We investigated what structured electronic health record data can predict about depression and anxiety progression in MS, and where its limits lie. We developed gradient boosting models to predict PHQ-9 (depression) and GAD-7 (anxiety) score change using EHR data from 2,163 MS patients (7,327 observations) and 1,465 patients (3,319 observations), respectively. Models achieved *R*^2^ of 0.22 (PHQ-9) and 0.28 (GAD-7). Baseline score was the dominant predictor, but this largely reflects regression to the mean: patients with high baseline scores tend to improve, while those with low scores tend to worsen. Age emerged as a consistent secondary predictor across both models: younger patients showed smaller improvements independent of baseline severity. Feature importance differed between models—PHQ-9 prediction relied on symptom subscales while GAD-7 incorporated pain and disease duration. These results suggest that structured clinical data alone capture only a fraction of what drives mental health trajectories, and that richer data sources—clinical notes, patient-reported outcomes, digital phenotyping—will be needed to enable meaningful individual-level prediction.

## 1 Introduction

In multiple sclerosis (MS), depression affects approximately 30.5% and anxiety 22% of patients based on pooled prevalence estimates, with rates of clinically significant symptoms reaching 35% and 34% respectively—substantially higher than the general population [1]. These conditions are frequently underdiagnosed and undertreated [2], and add substantially to disease morbidity and mortality, significantly impairing quality of life [3]. Validated screening instruments, including the Patient Health Questionnaire (PHQ-9) for depression [4] and Generalized Anxiety Disorder scale (GAD-7) for anxiety [5], are increasingly integrated into routine clinical care, generating longitudinal data that could enable proactive, personalized intervention.

Machine learning approaches have been applied to mental health prediction with mixed success. Machine learning approaches have achieved AUCs of 0.67–0.73 for cross-sectional depression and anxiety screening [6], while also enabling interpretability through methods like SHAP that reveal which features drive predictions [7]. Predicting future symptom severity or treatment response proves more challenging, with models achieving AUCs of 0.57– 0.77 [8, 9]. A consistent finding across studies is that baseline symptom severity dominates prediction— patients with high baseline scores tend to have higher future scores, though this relationship partly reflects regression to the mean and treatment patterns.

An intriguing finding from longitudinal MS research is that younger patients consistently report worse mental health than those diagnosed later in life, dePredicting Depression and Anxiety Progression in Multiple Sclerosis from Longitudinal Clinical Data Using Machine Learning spite having better physical health-related quality of life [10, 11, 12]. This may be explained by the theory of “off-time” life events [13]: younger individuals diagnosed with chronic illness face greater psychosocial disruption, as the disease intersects with life-stage demands such as establishing careers and relationships, while older adults demonstrate greater resilience relative to their peers. For MS specifically, the aetiology of depression is multifactorial, involving neurobiological mechanisms such as hippocampal microglial activation and regional atrophy, as well as the stressors and losses that accompany living with an unpredictable disease [14].

We developed machine learning models to predict PHQ-9 and GAD-7 score change in a large MS cohort using structured electronic health record (EHR) data. Our goals were to: (1) characterize what can be predicted from available clinical features, and (2) identify predictors of trajectory beyond regression to the mean. Our contributions are:

1. Development and validation of gradient boosting models for predicting depression and anxiety score change in MS patients
2. Characterization of baseline severity as the dominant predictor, reflecting regression to the mean, with age as a consistent secondary factor
3. Comparison of feature importance between depression and anxiety models, revealing distinct predictive profiles

## 2 Methods

### 2.1 Study Design

We conducted a retrospective cohort study of MS patients at an academic medical center with PHQ-9 or GAD-7 assessments in their electronic health records. Patients were included if they had a confirmed MS diagnosis and at least one valid baseline-follow-up assessment pair.

#### Temporal design

We defined baseline (*t*_0_) and follow-up (*t*_1_) timepoints for each observation (Figure 1b). For baseline selection, we identified all mental health assessments as candidate timepoints. When multiple candidates fell within 90 days, we selected the one with the fewest missing features. We chose 90 days because PHQ and GAD questionnaires assess symptoms over a 2-week recall period, and assessments closer together likely reflect overlapping symptom periods rather than independent observations. For follow-up selection, we identified the earliest subsequent assessment occurring 0.5–1.5 years after baseline. This window ensures sufficient time for clinically meaningful change while aligning with typical annual follow-up schedules. We avoided shorter intervals because more frequent assessments often indicate expected clinical changes (e.g., treatment initiation), introducing selection bias. A patient could contribute multiple non-overlapping *t*_0_–*t*_1_ pairs.

**Figure 1:**
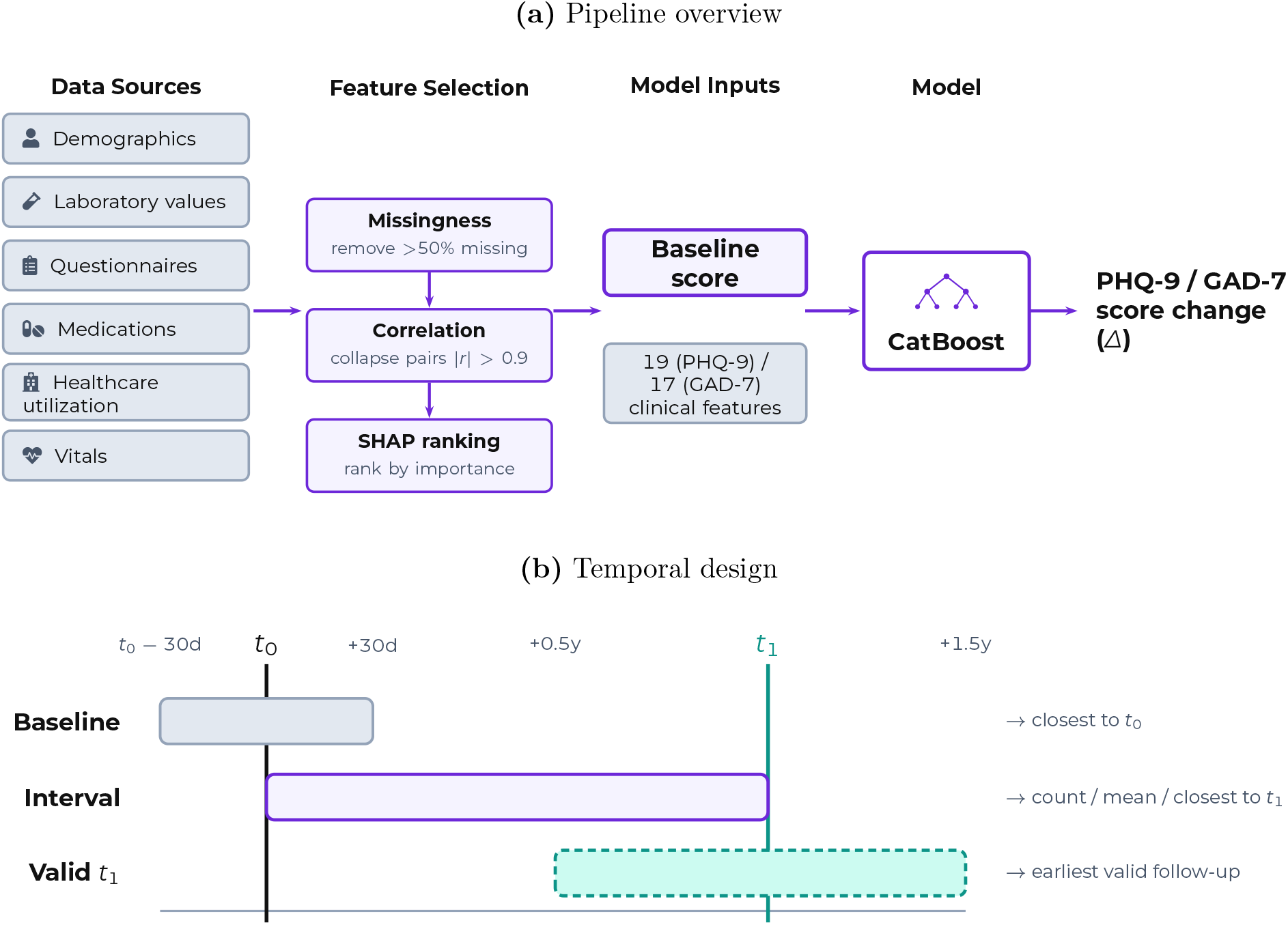
Study design. (a) Machine learning pipeline: candidate features are extracted from six categories of structured EHR data, filtered by missingness (*>*50%), correlation (*r >* 0.9), and SHAP importance. Final model inputs consist of the baseline mental health score—the dominant predictor—along with approximately 17 additional clinical features. (b) Temporal design for feature extraction: baseline features are extracted within ± 30 days of *t*_0_, selecting the value closest to *t*_0_. Interval features are collected between *t*_0_ and *t*_1_. The follow-up assessment (*t*_1_) is the earliest assessment 0.5–1.5 years after *t*_0_.

#### Outcome

The primary outcome was the score change between baseline and follow-up, defined as 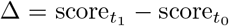. Positive values indicate worsening symptoms; negative values indicate improvement. Predicting score change rather than future score avoids inflated performance from autocorrelation between assessments. However, regression to the mean causes patients with extreme baseline scores to show predictable changes independent of clinical factors—high scores tend to improve, low scores tend to worsen.

### 2.2 Features and Preprocessing

We extracted candidate features from all structured data available in the institution’s research data warehouse, spanning demographics, laboratory values, clinical questionnaires, medications, healthcare utilization, and vitals (Figure 1a). From this pool of thousands of candidate features, we selected those with sufficient coverage and predictive value, as described below.

We organized features into two temporal categories (Figure 1b): baseline features from the ± 30 day window around *t*_0_, and interval features from the period between *t*_0_ and *t*_1_.

**Baseline features** capture the patient’s state at assessment: demographics (age, sex, race), MS disease characteristics (duration), medications (disease-modifying therapies, antidepressants, anxiolytics), healthcare utilization (visits, LACE+ score), laboratory values, and baseline mental health scores. For each feature, we selected the measurement closest in time to *t*_0_ within the ± 30 day window; if none existed, the feature was marked as missing.

**Interval features** capture events between assessments that may influence trajectory: rehabilitation (physical and occupational therapy), lifestyle factors (smoking, exercise), mental health treatment (therapy visits), and medication use. These features likely serve as proxies for healthcare engagement or care intensity rather than direct causal factors. For features with multiple occurrences, we computed counts, means, or selected the value closest to *t*_1_.

#### Feature selection

We first removed features with *>*50% missingness and highly correlated pairs (Pearson *r >* 0.9), retaining the feature with lower missingness. We excluded observations where *>*50% of remaining features were missing. We then ranked remaining features by SHAP importance and evaluated model performance as features were added. Performance plateaued at approximately 20 features for both models, with additional features providing negligible improvement.

### 2.3 Statistical Analysis

We used CatBoost gradient boosting [15], selected for its native handling of categorical features and compatibility with SHAP-based interpretability [16].

#### Training and validation

We split data 80/20 at the patient level—all observations from a given patient were assigned exclusively to training or test set, preventing data leakage from repeated measurements [17]. Hyperparameters were tuned using Optuna [18] on a held-out validation set (20% of training data, split at the patient level), optimizing for RMSE. Final models were evaluated using standard regression metrics (*R*^2^, MAE, RMSE) on the held-out test set. The hyperparameter search space included: tree depth (1–12), learning rate (0.001–0.3), L2 regularization (1–10), bootstrap type (Bayesian, Bernoulli, or MVS), random strength (0–10), and feature fraction per tree level (0.01–0.1). The number of iterations was fixed at 500.

#### Evaluation

We assessed model performance on the held-out test set using *R*^2^ (variance explained), mean absolute error (MAE), and root mean squared error (RMSE). Feature importance was quantified using SHAP values [7], with beeswarm plots showing how feature values affect predictions.

#### Data aggregation

To comply with institutional data sharing requirements that protect patient privacy, SHAP values were exported in aggregated form. For each feature, patients were sorted by their SHAP values and grouped into bins of exactly 10 patients. We then computed the mean SHAP value and mean feature value for each bin. This aggregation prevents identification of individual patients while preserving the relationship between feature values and their effects on predictions—each point in our beeswarm plots represents one such bin rather than an individual patient.

## 3 Results

### 3.1 Cohort Characteristics

The source population included 18,252 MS patients from the medical center (Figure 2). The population was predominantly female (72%) and white (88%), with a mean age of 54.9 years (SD 15.2). Most patients were in the 40–70 age range, consistent with a prevalent MS cohort given a typical age of diagnosis around 32 years [19]. The cohort had extensive longitudinal follow-up: 55% of patients had at least 5 years of data, 37% had 10 years, and 13% had 20 or more years. Most patients were seen regularly, with a median interval of 33 days between visits; the majority attended monthly or bi-weekly appointments.

**Figure 2:**
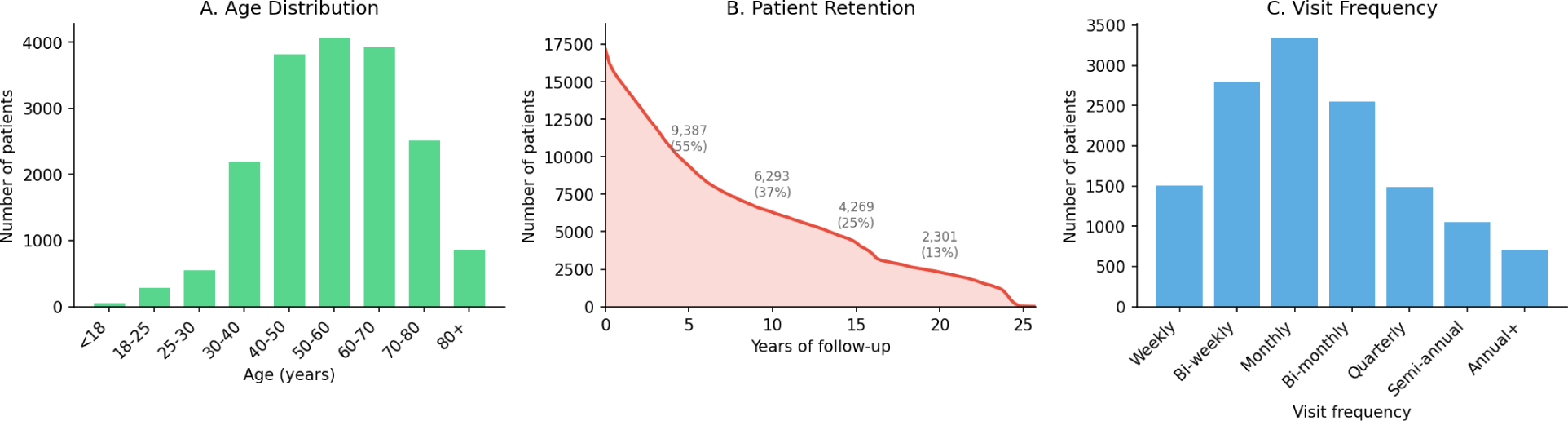
Source population characteristics. (A) Age distribution of the 18,252 MS patients at the medical center. (B) Patient retention over time, showing counts and percentages at 5, 10, 15, and 20 years of followup. (C) Distribution of visit frequency patterns. Subsets of 2,163 (PHQ-9) and 1,465 (GAD-7) patients with valid assessment pairs were included in the prediction analysis.

Of these, 2,163 patients contributed 7,327 valid PHQ-9 baseline-followup pairs and 1,465 patients contributed 3,319 valid GAD-7 pairs meeting inclusion criteria (0.5–1.5 year interval). The smaller GAD-7 cohort reflects later adoption of anxiety screening in routine clinical workflows.

### 3.2 Prediction Performance

Model performance was modest (Figure 3). For PHQ-9, *R*^2^ = 0.22, MAE = 3.5, RMSE = 4.7. For GAD-7, *R*^2^ = 0.28, MAE = 3.2, RMSE = 4.4. The models explain 22–28% of variance in score change; the remaining variance reflects factors not captured in structured EHR data, including life events, social support, and therapeutic engagement.

**Figure 3:**
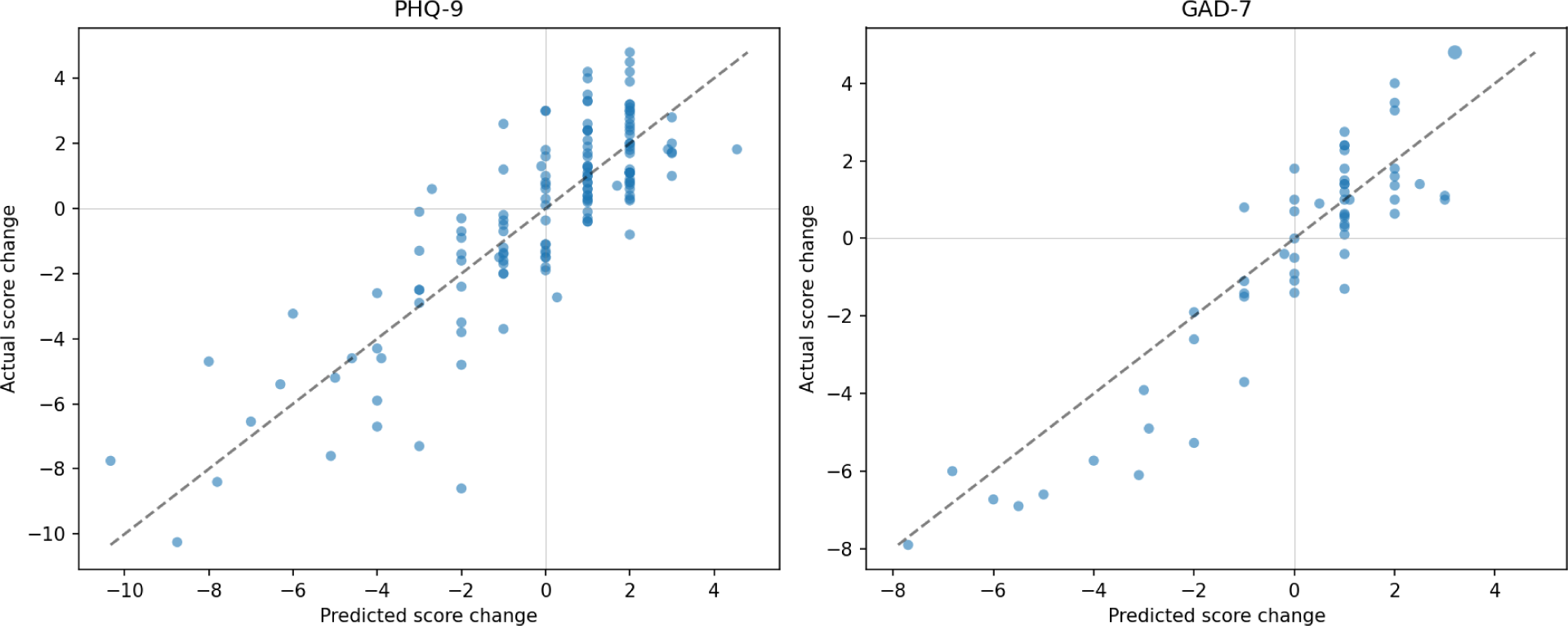
Predicted vs. actual score change for PHQ-9 (left) and GAD-7 (right). A dashed line indicates perfect prediction. The models explain 22–28% of variances in score change.

**Figure 4:**
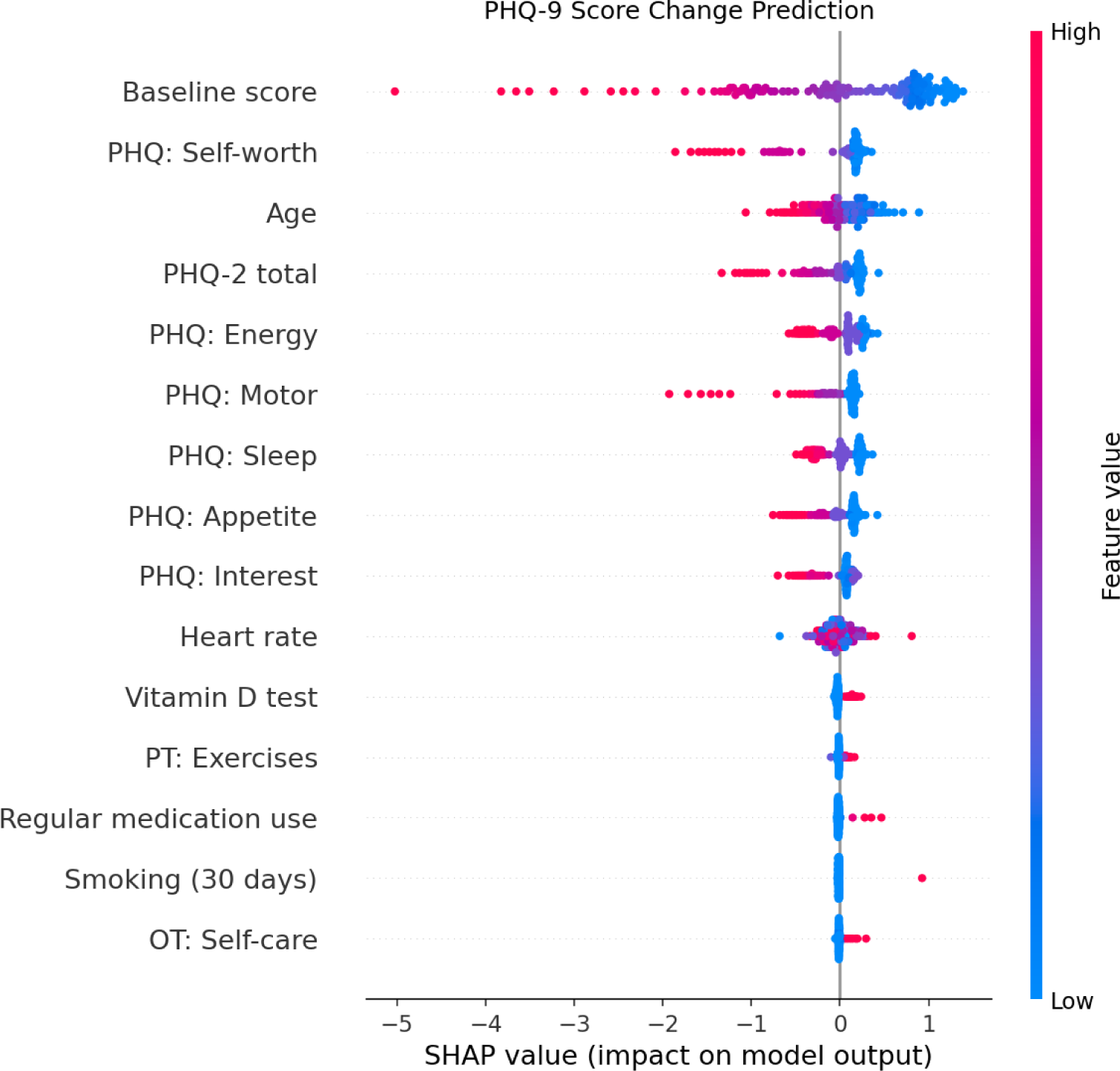
SHAP feature importance for PHQ-9 score change prediction. Each point represents one aggregated bin (minimum 10 patients per bin for privacy). Color indicates feature value (blue=low, red=high); horizontal position shows impact on predicted score change. High baseline scores predict negative change (improvement), reflecting regression to the mean.

**Figure 5:**
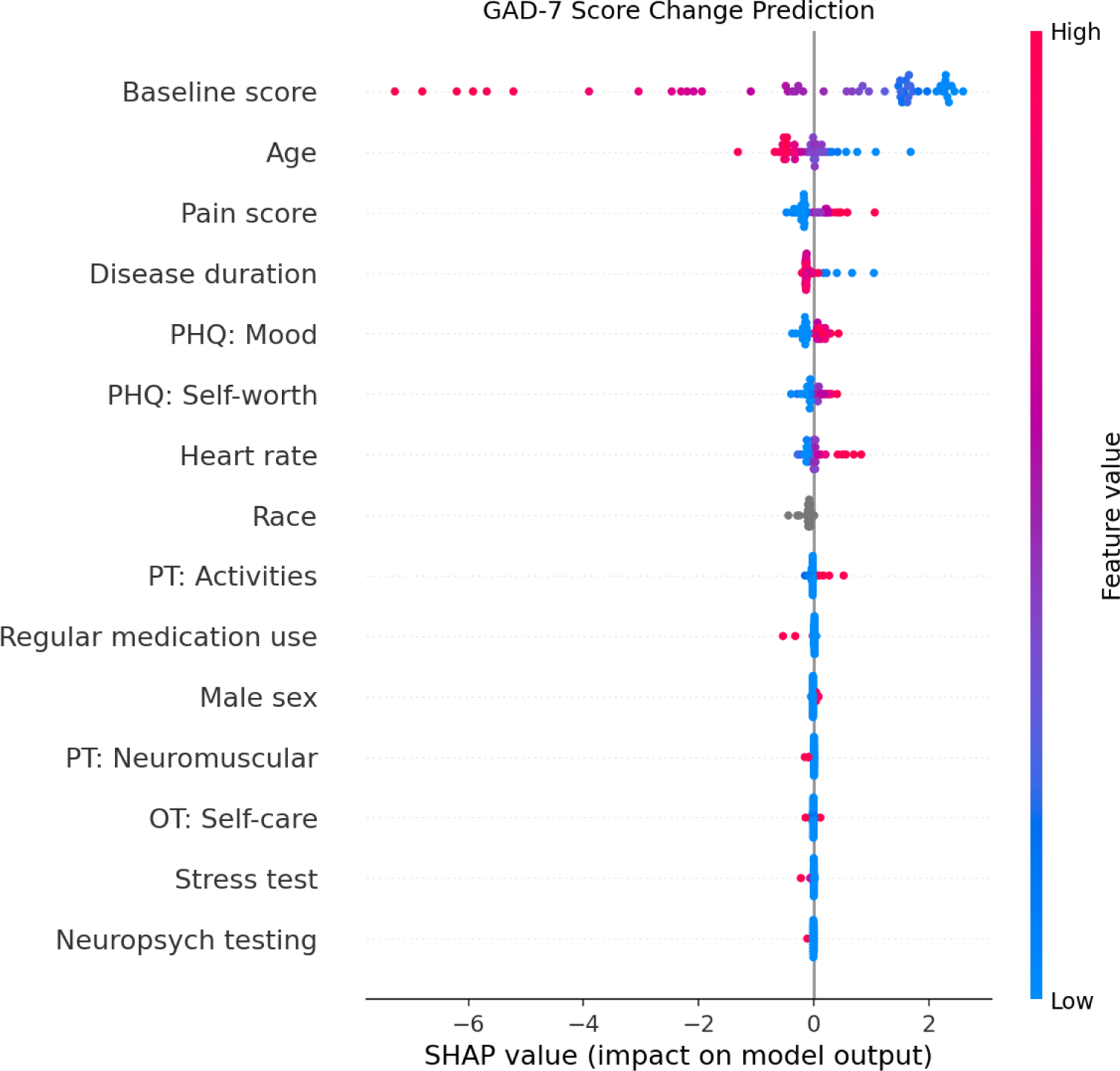
SHAP feature importance for GAD-7 score change prediction. Baseline score, age, and pain score are the dominant predictors. Younger patients (blue) show positive SHAP values, indicating smaller improvements or greater worsening.

### 3.3 Feature Importance

#### 3.3.1 Baseline Score and Regression to the Mean

Baseline questionnaire score was by far the most important predictor (mean |SHAP| = 0.97 for PHQ-9, 2.16 for GAD-7). However, the direction of this effect reflects regression to the mean: higher baseline scores predicted negative change (improvement), while lower baseline scores predicted positive change (worsening). For PHQ-9 prediction, patients with high baseline scores (PHQ-9 ∼ 25) had mean SHAP values of − 5.0 (predicting improvement), while those with low baseline scores had positive SHAP values (predicting worsening). This statistical phenomenon—where extreme values tend to move toward the population average—partially explains why baseline severity dominates prediction of change.

To quantify the extent of this dominance, we decomposed total SHAP attribution across all features. Baseline score accounted for 31% of a total mean |SHAP| in the PHQ-9 model and 59% in the GAD-7 model. The remaining 69% (PHQ-9) and 41% (GAD-7) of predictive attribution came from clinical features beyond baseline—indicating that while regression to the mean is substantial, structured EHR data does capture additional trajectory signal. The contrast between models is notable: PHQ-9 prediction draws the majority of its signal from symptom subscales and clinical features, whereas GAD-7 prediction relies more heavily on baseline severity alone.

#### 3.3.2 Age Effect

Beyond baseline score, age emerged as a consistent secondary predictor (mean SHAP = 0.28 for PHQ-9, 0.38 for GAD-7). Younger patients showed positive |SHAP| values (predicting smaller improvements or greater worsening), while older patients showed negative SHAP values (predicting greater improvements). This represents approximately a 1–2 point difference in predicted score change across the age range, independent of baseline severity.

#### 3.3.3 Model-Specific Features

Feature importance differed notably between depression and anxiety models.

**PHQ-9 prediction** relied heavily on symptom subscales beyond the total score. Self-worth (mean |SHAP| = 0.31), energy (0.23), motor symptoms (0.23), and sleep (0.20) all contributed independently. This suggests that symptom profile—not just total severity—informs trajectory prediction.

**GAD-7 prediction** incorporated clinical factors beyond mental health symptoms. Pain score (mean |SHAP| = 0.26) and disease duration (0.18) emerged as predictors, consistent with the established link between chronic pain, illness burden, and anxiety in MS. Heart rate (0.14) and PHQ mood item (0.16) also contributed, suggesting that physiological arousal and comorbid depression influence anxiety trajectories.

## 4 Discussion

### 4.1 Key Findings

Our analysis reveals both the potential and limitations of predicting mental health trajectories in MS.

#### Prediction of change is challenging

The models explained 22–28% of variance in score change. Predicting change rather than future score removes the autocorrelation between consecutive assessments, making the task inherently harder. The remaining variance reflects factors not captured in structured clinical data: life events, social support, coping mechanisms, and therapeutic engagement.

#### Regression to the mean is substantial but not the whole story

Baseline score was the strongest single predictor of change, and its direction—high scores predicting improvement, low scores predicting worsening—reflects regression to the mean. However, SHAP decomposition reveals that baseline accounts for only 31% (PHQ-9) to 59% (GAD-7) of total predictive attribution. The remaining signal comes from clinical features: symptom subscales, age, pain, and disease duration. This decomposition quantifies what is often assumed but rarely measured—the relative contribution of statistical artifact versus clinical signal in longitudinal prediction.

#### Age is a consistent secondary predictor

Younger patients showed smaller improvements (or greater worsening) independent of baseline severity. This aligns with findings from chronic illness research showing that older adults demonstrate greater psychosocial resilience relative to their peers than younger adults do [13]. For younger patients, chronic illness represents an “off-time” disruption to expected developmental milestones [13, 10].

#### Feature importance differs between models

PHQ-9 prediction relied on symptom subscales, suggesting that symptom profile informs trajectory beyond total severity. GAD-7 prediction incorporated pain and disease duration, consistent with the link between chronic pain and anxiety in MS.

### 4.2 Clinical Implications

Despite modest predictive accuracy, our findings have practical implications:

#### 1. Targeted monitoring

Younger MS patients may warrant more frequent mental health screening, independent of current symptom level

#### 2. Symptom persistence

Patients with elevated scores should be counseled that symptoms tend to persist without intervention, emphasizing the importance of early treatment. Evidence-based interventions include structured exercise programs, which show moderate effects on depression across populations (median standardized mean difference −0.43) [20]

#### 3. Prediction vs. intervention

While our models identify risk factors, the dominant predictors (baseline severity, age) are not modifiable. Clinical value lies in identifying patients who need intervention, not in discovering new intervention targets

### 4.3 Limitations

#### Regression to the mean limits interpretability

As quantified in our SHAP decomposition, baseline score accounts for 31–59% of predictive attribution. Because baseline score enters the change-score outcome with a negative sign, this dominance partly reflects regression to the mean and mathematical coupling between a change and its own baseline rather than a clinical mechanism. The genuinely predictive, clinically actionable signal is therefore confined to the remaining attribution carried by symptom subscales, age, pain, and disease duration.

#### Unmeasured confounders

Key determinants of mental health (social support, life stressors, therapy engagement) are not captured in structured clinical data.

#### Sparse data

Despite filtering, remaining feature missingness may affect model performance and feature importance rankings.

#### Observational design

We cannot infer causality; age and other identified features are associations, not validated intervention targets.

### 4.4 Future Directions

Several approaches could improve mental health trajectory prediction. First, aggregating sparse but related features may recover signal lost to missingness filtering. Individual rehabilitation features (physical therapy, occupational therapy, speech therapy) are often too sparse to retain, but a combined “rehabilitation intensity” measure might capture meaningful treatment engagement that predicts improvement.

Second, clinical notes contain rich information not reflected in structured data—patient-expressed concerns, social circumstances, clinician observations about affect and coping. NLP can identify mental illness documentation in patients whose structured records lack such information [21], and could extract predictive signals such as mentions of life stressors, hopelessness, or social isolation that structured EHR fields cannot capture.

Third, and perhaps most promising, is the integration of patient-reported outcomes collected outside clinical encounters. Current EHR data captures what clinicians do (prescriptions, referrals) but not what patients experience between visits. Smartphone-based ecological momentary assessments could track mood, sleep, and activity at higher frequency than clinical visits [22]. Recent work in MS specifically has shown that combining passive sensing from smartphones and fitness trackers with brief self-reported mood and fatigue questions can predict depressive symptoms with 80.6% accuracy [23], and EMA-derived features achieved substantially higher predictive accuracy for next-day suicidal ideation than passive sensor data alone [24], suggesting that subjective patient experience remains essential for prediction. This shift from clinic-centric to patient-centric data collection may be necessary to substantially improve the prediction of mental health trajectories.

## Conclusion

Predicting mental health score change in MS from structured clinical data is challenging, with models explaining 22–28% of variance. Baseline severity was the dominant predictor, primarily reflecting regression to the mean—patients with high scores tend to improve, those with low scores tend to worsen. Age emerged as a consistent secondary predictor, with younger patients showing smaller improvements independent of baseline. Feature importance differed between depression and anxiety models: PHQ-9 prediction relied on symptom subscales while GAD-7 incorporated pain and disease duration. These findings highlight both the statistical challenges and clinical opportunities in EHR-based trajectory prediction for mental health in MS.

## Acknowledgement

The authors would like to thank the Ministry of the Economy in Luxembourg and its digital health directorate for co-supporting this research. Similarly, we would like to thank the Luxembourg National Research Fund (FNR) for partially funding this research.

## Data Availability

Due to patient privacy requirements and institutional data governance policies, the individual-level data used in this study cannot be shared publicly. Aggregated SHAP values and summary statistics are available upon reasonable request to the corresponding author.

## A Feature Descriptions

This appendix provides detailed descriptions of all features used in the prediction models. Features are categorized by type and listed in order of SHAP importance (mean |SHAP| value).

### A.1 PHQ-9 Model Features

**Table 1:** Features used in PHQ-9 score change prediction model, ordered by SHAP importance.

| Feature | Category | Description | SHAP |
| --- | --- | --- | --- |
| Baseline score | Mental health | PHQ-9 total score at $t_0$ (0–27) | 0.97 |
| PHQ: Self-worth | Mental health | “Feeling bad about yourself” item (0–3) | 0.31 |
| Age | Demographics | Age in years at baseline assessment | 0.28 |
| PHQ-2 total | Mental health | PHQ-2 screening score at $t_0$ (0–6) | 0.24 |
| PHQ: Energy | Mental health | “Feeling tired or having little energy” item (0–3) | 0.23 |
| PHQ: Motor | Mental health | “Moving or speaking slowly/being fidgety” item (0–3) | 0.23 |
| PHQ: Sleep | Mental health | “Trouble falling or staying asleep” item (0–3) | 0.20 |
| PHQ: Appetite | Mental health | “Poor appetite or overeating” item (0–3) | 0.19 |
| PHQ: Interest | Mental health | “Little interest or pleasure in doing things” item (0–3) | 0.15 |
| Heart rate | Clinical | Resting heart rate in bpm, closest to $t_0$ | 0.13 |
| Vitamin D test | Interval | Any 25-hydroxyvitamin D lab test between $t_0$–$t_1$ (binary) | 0.05 |
| Regular medication use | Interval | Reports regular use of medications, vitamins, or supplements (binary) | 0.03 |
| PT: Exercises | Interval | Physical therapy exercise session between $t_0$–$t_1$ (binary) | 0.03 |
| Smoking (30 days) | Interval | Reported smoking in last 30 days between $t_0$–$t_1$ (binary) | 0.03 |
| OT: Self-care | Interval | Occupational therapy self-care training between $t_0$–$t_1$ (binary) | 0.02 |
| PT: Activities | Interval | Physical therapy activities session between $t_0$–$t_1$ (binary) | 0.01 |
| PT: Neuromuscular | Interval | Neuromuscular reeducation session between $t_0$–$t_1$ (binary) | 0.01 |
| Stress echo | Interval | Stress echocardiogram between $t_0$–$t_1$ (binary) | 0.01 |
| Neuropsych testing | Interval | Neuropsychological evaluation between $t_0$–$t_1$ (binary) | 0.01 |
| Current smoker | Interval | Currently uses tobacco products (binary) | < 0.01 |

### A.2 GAD-7 Model Features

**Table 2:**
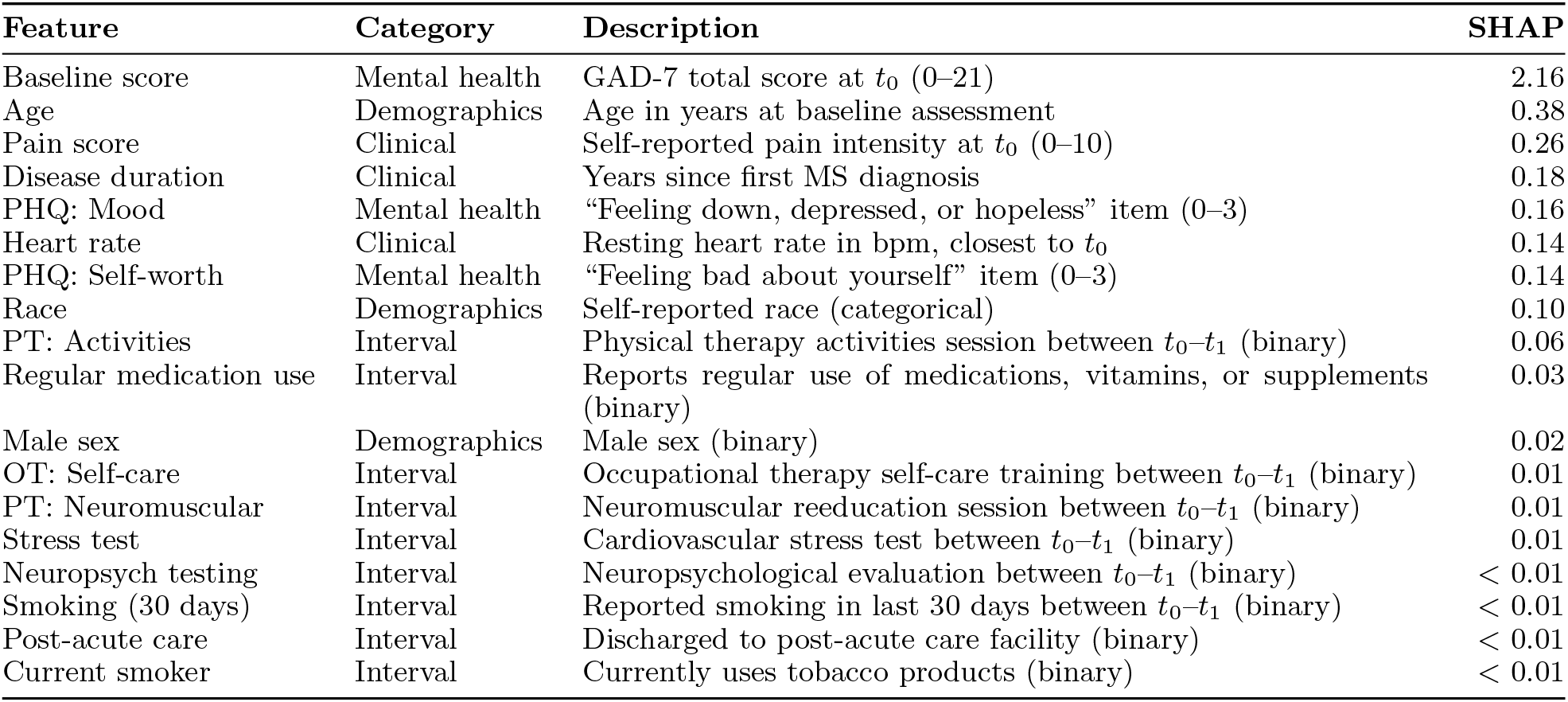
Features used in GAD-7 score change prediction model, ordered by SHAP importance.

